# Toward a preventive approach to prolonged grief disorder in palliative care: Insecure attachment moderates the impact of perceived support on the severity of symptoms

**DOI:** 10.1101/2022.03.18.22272629

**Authors:** Vittorio Lenzo, Alberto Sardella, Cristina Faraone, Maria C. Quattropani

**Author notes:** Correspondence concerning this article should be addressed to Vittorio Lenzo, Department of Social and Educational Sciences of the Mediterranean Area, University for Foreigners “Dante Alighieri”, Via del Torrione 95, 89125, Reggio Calabria, Italy.

## Abstract

**Objective:** This study aimed to investigate the relationships between the perceived support at the time of assistance, insecure attachment (i.e., avoidance and anxious attachment factors), and the prolonged grief symptoms in family caregivers of palliative care patients deceased for at least one year. We also investigated the moderating role of insecure attachment in the relationship between perceived support and intensity of prolonged grief symptoms.

**Method:** A sample of 157 participants completed the Prolonged Grief Scale (PG-13) and the Attachment Style Questionnaire (ASQ).

**Results:** Correlational analysis indicated that prolonged grief symptoms were positively correlated with the Avoidance attachment factor but not with the Anxious one. The perceived support was negatively related to both the Avoidance and the Anxious attachment factors. Lastly, the two insecure attachment factors were moderately and positively correlated with each other. Results of moderation analysis showed that the high Avoidance attachment moderated the effect of the perceived family and social support on the intensity of prolonged grief symptoms among family caregivers of patients assisted in palliative home care. Results also showed that the Anxious attachment factor had a significant effect on prolonged grief symptoms, even though the interaction with the perceived support was not significant.

**Conclusions:** Overall, these results underline that a high level of avoidance attachment may moderate the relationship between the perceived support and intensity of grief symptoms and thereby increasing the risk of developing a mental disorder. Intervention to prevent prolonged grief disorder among family caregivers should take into account these findings.

## Introduction

In Italy, recent estimates reported more than 180.000 deaths for cancer [1] and almost 40.000 received palliative home care [2]. When supporting their suffering loved ones, family caregivers of patients with advanced cancer face a stressful event that may have mental health consequences even after the loss [3]. Most of the bereaved regain quickly a normal functioning, while others may experience acute symptoms at the beginning [4]. Nonetheless, a pooled prevalence of 9.8 percent show symptoms of an abnormal grief reaction categorized as prolonged grief disorder (PGD) [5]. Proposed criteria for PGD included separation distress, together with cognitive, emotional, and behavioral symptoms [6]. Furthermore, the symptoms that characterize PGD are elevated at least 6 months after the loss and cause significant functional impairment. Prevention of prolonged grief disorder among bereaved is paramount. A central tenet of palliative care is to provide support during the patient’s illness [7], even though scant attention has been paid to what happens after the death of the loved one. Such an approach is of benefit in avoiding the onset of the disorder and claiming the role of health psychology in its prevention [8]. If only a part of bereaved individuals shows an abnormal grief reaction, understanding the role of specific factors increasing the risk for prolonged grief disorder is challenging, even though fundamental for prevention. So far, researchers within the field have repeatedly highlighted the gap between the policy and practice for bereavement support and prevention of prolonged grief disorder in the context of palliative care [9]. Worth noting, since the promising implications for implementation of the preventive approach, several studies have found significant relationships between complicated grief and a broad array of socio-demographic and psychological variables, including social support and insecure attachment style [10-12]. However, because of the limitations inherent to grief research, there be left many unanswered questions about the interplay between the potential predictor variables. For example, research based on data from the Changing Lives of Older Couples (CLOC) study examining the moderator role of social support in the relationship between bereavement and distress levels has found no significant effect [13-14]. On the other hand, another study, focusing on the pre-loss predictors of grief reaction patterns, found that chronic grievers reported less instrumental support, but not social support than resilient individuals [15]. Whatever the cause of these findings, it is reasonable to hypothesize that personality variables may play a moderator role in the relationship between the perceived social support during the assistance and the bereavement outcome. Turning to the psychological predictors, attachment theory has been considered a useful tool for understanding grief reaction [16]. Bowlby considered attachment as an innate motivational system that leads the individuals, from the cradle to the grave, to seek proximity with beloved figures in order to feel safe and secure [17]. According to attachment theory, positive childhood experiences with the caregivers are critical for developing secure internal working models of relationships and adequate emotion regulation abilities [18]. In the last decades, several studies have linked the quality of early attachment experiences with attachment relationships in adulthood [19-20]. Perhaps not surprisingly then, people who did not attain a secure attachment insofar their interactions with attachment figures during childhood were gone awry, have a higher likelihood of developing a mental disorder during adulthood [21]. In this vein, attachment research during adulthood has relied on the recurrent pattern of expectancy, feelings, and behavior characterizing the relationships of the individuals and resulting from their history of attachment experiences. The burgeoning literature on the person’s attachment style has focused attention on two relatively independent dimensions of insecure attachment named attachment-related anxiety and attachment-related avoidance and analogous to those observed in children [22-23]. Whereas individuals with avoidant attachment tend to preserve emotional independence from adult attachment figures and to distrust their goodwill, individuals with an anxious attachment tend to worry that attachment figures will be inaccessible or elusive in times of distress [20]. The loss of a loved one represents a potentially overwhelming that strongly strikes the attachment system and thereby can undermine the sense of security of the mourner [16]. Although feelings elicited by grief may vary hugely among bereaved individuals [24], its resolution hinges on the secondary attachment strategies of hyperactivation and deactivation [25]. These strategies are tantamount to the avoidance and anxious factors and virtually most of the studies have found a relationship with grief reactions [10]. To date, however, grief research leaves unsolved several crucial questions. A major research question regards the role of attachment in adjusting to the loss of a loved one. While attachment theory has posited that the lack of distress after the loss characterizing avoidant attachment style may lead to difficulties in the long run, it turned out that multiple pathways are possible, including seemingly maladaptive reactions [26]. Unfortunately, despite the relevance of understanding grief reactions, and perhaps due to the difficulties in conducting research in this area, very few studies have investigated the interplay between insecure attachment factors and prolonged grief in the context of palliative home care. For example, in a study involving a sample of 60 family caregivers of cancer patients assisted in hospice, Lai and colleagues found that preoccupation with relationships, which is a dimension of the high order factor of anxious attachment, contributed to predict the prolonged grief risk [27].

Although it has been hypothesized that insecure attachment style may have a moderator effect, changing how family and social support is perceived [28], there is still a lack of studies in the context of grief reactions in palliative home care. Based on these premises, the present study has two main objectives. First, we sought to examine the relationships between the perceived family and social support at the time of assistance, attachment insecurities (i.e., avoidance and anxious attachment factors), and the prolonged grief symptoms in a sample of family caregivers of palliative care patients deceased for at least one year. We hypothesized that avoidance and anxious attachment factors were positively correlated with each other and that, which in turn, were positively correlated with prolonged grief symptoms and negatively with the perceived family and social support. Second, we sought to investigate the moderating role of avoidance and anxious attachment factors in the relationship between perceived family and social support at the time of assistance and grief intensity in family caregivers of palliative home care patients. The conceptual models representing the investigated variables in a moderate relationship are illustrated in Figure 1. We hypothesized that avoidance and insecure attachment factors are moderators of the relationship between the perceived support and intensity of grief symptoms.

**Figure 1.**
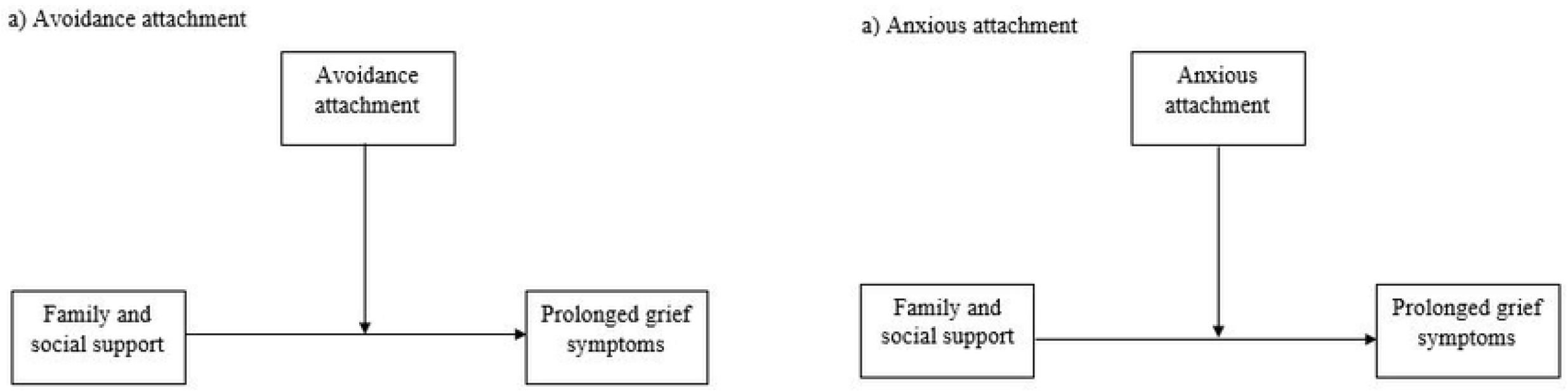
Conceptual model of the moderated role of Avoidance (a) and Anxious (b) attachment factors in the relationship between family/social support and prolonged grief symptoms

## Materials and Methods

### Participants and Procedure

This study is part of a larger research project named “Risk and protective factors for prolonged grief disorder in family caregivers of patients in palliative home care”. Participants were recruited between 7 February 2020 and 9 November 2021 among the caregivers of cancer patients who were assisted in three palliative home care services of three relatively small cities (Agrigento, Caltanissetta, and Messina) in Sicily, Italy. The inclusion criteria were: being at least 18 years old, able to give informed consent, and having lost for cancer a family member at least one year before enrolling in the study. Exclusion criteria were a pre-existing mental health disorder and/or taking psychotropic drugs. All participants completed the survey anonymously and gave informed consent before participating. Privacy of the participants was guaranteed in accordance with the European Union General Data Protection Regulation 2016/679. The research was conducted in accordance with the 1964 Declaration of Helsinki and its later amendments. This study was approved by the Research Ethics Committee for Psychological Research of the University of Messina (n. 38512). One hundred and fifty-nine subjects consented to participate and 157 completed the protocol.

### Measures

The participants completed a questionnaire including single-item questions on socio-demographic (i.e., age, gender, and education), loss (i.e., year of bereavement, relationship with the patient, and work status), and family and social support variables. Regarding the family and social support, the participants were asked to respond to the following question: “How much did you feel supported by your family and friends in the time of palliative care?” The response was rated on a 10-point Liker scale from 1 (“for nothing”) to 10 (“very much”).

Moreover, the following two self-report measures were administered:

The Prolonged Grief Scale (PG-13) [29] is a self-report instrument to assess grief intensity and to diagnose the prolonged grief disorder related to the DSM-5 [6] and ICD-11 criteria [30]. The PG-13 consists of 11 items on a five-point Likert scale which assesses the severity/intensity of prolonged grief disorder. Indeed, each item refers to a symptom of prolonged grief disorder, such as cognitive, emotional, and behavioral symptoms. A total score consisting of the sum of all the items can be computed to obtain the intensity of the symptoms. Two more items are rated “yes”/no” and permit to examine if the symptoms of separation distress are experienced more than once a day and if are related to a significant diminution of social and occupational functioning. In the current study, the Italian version of PG-13 [31] showing adequate psychometric properties was used. The degree of reliability for this sample was excellent, with a Cronbach’s α of 0.88.

The Attachment Style Questionnaire (ASQ) [32] is a self-report instrument developed to evaluate attachment style factors. The 40 items of the questionnaire are scored on a six-point Likert scale which examines the following dimensions: Confidence (C); Discomfort with closeness (DwC); Relationship as secondary (RaS); Need for approval (NfA); Preoccupation with relationship (PwR). Following Hazan and Shaver’s [20] and Bartholomew’s [33] definition of attachment styles, the authors of ASQ endorsed the insecure anxious/ambivalent attachment (through NfA and PwR) and insecure-avoidant attachment (through DwC and RaS). In the present study, the Italian version of ASQ [34] showing acceptable psychometric properties was adopted. For the purpose of this study, the two-factor higher order structure of the ASQ, consisting of Avoidance and Anxious attachment factors, was used. The degree of reliability for this sample was excellent, with a Cronbach’s α ranging from 0.65 for Confidence to 0.76 for Relationship as secondary.

### Statistical Analysis

The data were analyzed using SPSS v. 26 (IBM, Armonk, NY, USA) statistical software and the Process Macro for SPSS [35]. Descriptive results were described as frequencies (%), mean scores, and standard deviations. Relationships between perceived support, PG-13, and ASQ-40 were performed with Pearson product-moment correlation coefficients. Subsequently, two distinct moderation analyses were performed to examine the role of the Avoidance and Anxious attachment factors in moderating the effect of family and social support on the PG-13. Specifically, we considered the perceived support as the independent variable, the PG-13 as the dependent variable, and the Avoidance attachment factor and the Anxious attachment factor as the moderator. In each moderation analysis, the effect of interaction (family and social support x attachment insecurities factors) was decomposed through simple slope analysis at low (−1 *SD*), medium, and high (+1 *SD*) values of the moderator. We also used the Johnson-Neyman method to identify the exact value of the moderator at which the effect became significant. All the variables that define the product were centered, even though we choose to indicate raw values for the Johnson-Neyman solution and the figure for better clarity in understanding the results.

## Results

### Socio-demographic and loss characteristics of the sample

The socio-demographic and loss characteristics of the sample are shown in Table 1. The final sample consisted of 157 subjects who ranged in age from 18 to 81 years (*M* = 43.50 *±* 14.04). Most of the participants were female (77.1*%*), had a high school diploma (42*%*), and worked after loss (59.87*%*). Among participants, 52.3*%* were sons or daughters to the deceased and 57.3*%* were the main caregiver, while the time since the loss was on average 3.59 years (*SD* = 4.92).

**Table 1.**
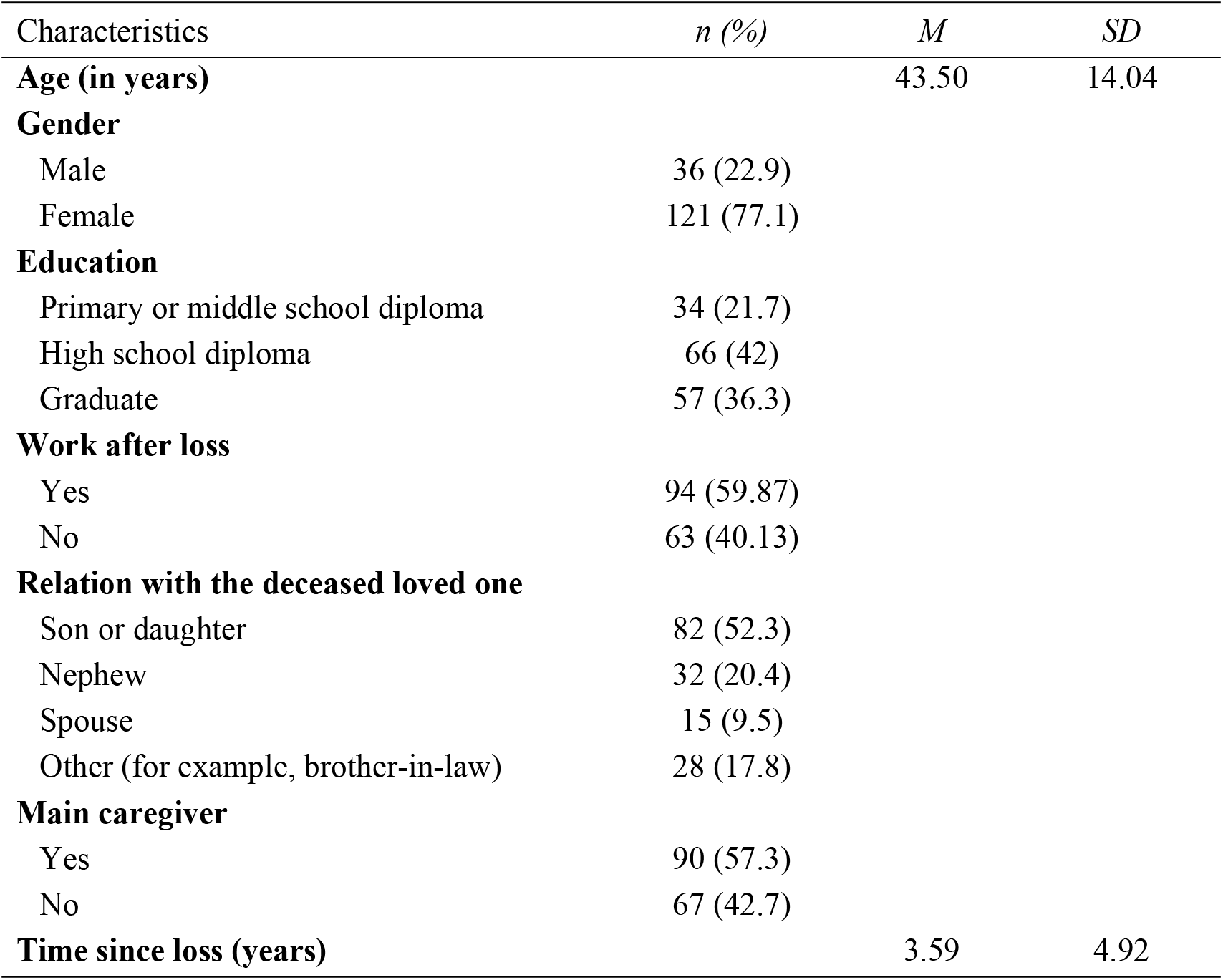
Socio-demographic and loss characteristics of the sample (N = 157)

### Correlational analysis between family and social support, prolonged grief symptoms intensity, and attachment style factors

Table 2 displays descriptive statistics and correlation analyses. Results showed that family and social support was negatively and weakly correlated with both the Avoidance attachment factor (*r* = -.17; *p* < .05) and the Anxious attachment factor (*r* = -.25; *p* < .01). Results also showed that PG-13 was positively and weakly correlated with the Avoidance attachment factor (*r* = .25; *p* < .01)) but not with the Anxious attachment factor and family/social support. Lastly, the two attachment insecurities factors were moderately and positively correlated (*r* = .43; *p* < .01).

**Table 2.** Descriptive and correlational analysis (*N* = 157)

| Variable | *Min* | *Max* | *M* | *SD* | *Skew* | *Kurt* | 1 | 2 | 3 |
| --- | --- | --- | --- | --- | --- | --- | --- | --- | --- |
| 1. Family/social support | 1 | 10 | 7.73 | 2.34 | -1.17 | 1.04 | - |  |  |
| 2. Prolonged Grief Scale – 13 | 11 | 53 | 27.08 | 8.84 | 0.52 | -0.16 | .08 | - |  |
| 3. Avoidance attachment factor | 32 | 93 | 56.31 | 11.22 | 0.32 | 0.19 | -.17* | .25** | - |
| 4. Anxious attachment factor | 19 | 81 | 47.03 | 12.68 | 0.25 | -0.35 | -.25** | .18 | .43** |

*Note.* *Min* minimum value, *Max* maximum value, *M* mean, *SD* standard deviation, *Skew* skewness, *Kurt* kurtosis
\* $p < .05$ ; \*\* $p < .01$ .

### Moderation of the family and social support – prolonged grief symptoms association by the attachment insecurities factors

Two separate moderation models were carried out to test whether the Avoidance and the Anxious attachment factors moderated the effect of the perceived family and social support at the time of assistance on the intensity of prolonged grief symptoms.

Table 3 shows that the Avoidance attachment factor (but not the perceived support) had a significant effect on the prolonged grief symptoms (*B* = 0.23; *p <* .01). Moreover, the Family and social support predicted the severity of prolonged grief symptoms only at high levels of the Avoidance attachment factor (*B* = 0.05; *p <* .05), but not at low or medium ones. Figure 2 represents the different slopes for the conditional effect of the perceived family and social support on the prolonged grief symptoms highlighting the role of the Avoidance attachment factor. The Johnson-Neyman method showed that for caregivers with the Avoidance attachment score lower than 7.94, family and social support was no longer related to the severity of prolonged grief symptoms.

**Table 3.** Avoidance attachment moderates the relationship between Family/social support and intensity of prolonged grief symptoms

|  | *B* | *SE* | *t* | 95% CI |
| --- | --- | --- | --- | --- |
| Intercept | 27.28 | 0.68 | 39.00 | [25.93, 28.63] |
| Family/social support | 0.23 | 0.31 | 0.74 | [-0.39, 0.85] |
| Avoidance attachment | 0.23 | 0.06 | 3.72** | [0.11, 0.36] |
| Family/social support x<br>Avoidance attachment | 0.05 | 0.02 | 2.03* | [0.01, 0.09] |
| Regression Model $R^2 = .10^{**}$ | | | | |
| $\Delta R^2 = .02^*$ | | | | |
| Conditional effects of<br>Family/social support |  |  |  |  |
| Low Avoidance attachment (-1SD<br>= -11.22) | -0.28 | 0.47 | -0.61 | [-1.21, 0.64] |
| Medium Avoidance attachment<br>($M = 0$) | 0.23 | 0.31 | 0.74 | [-0.39, 0.85] |
| High Avoidance attachment<br>(+1SD = 11.22) | 0.75 | 0.33 | 2.28* | [0.10, 1.40] |

Note. CI = confidence interval.
\* $p < .05$ ; \*\* $p < .01$ .

**Figure 2.**
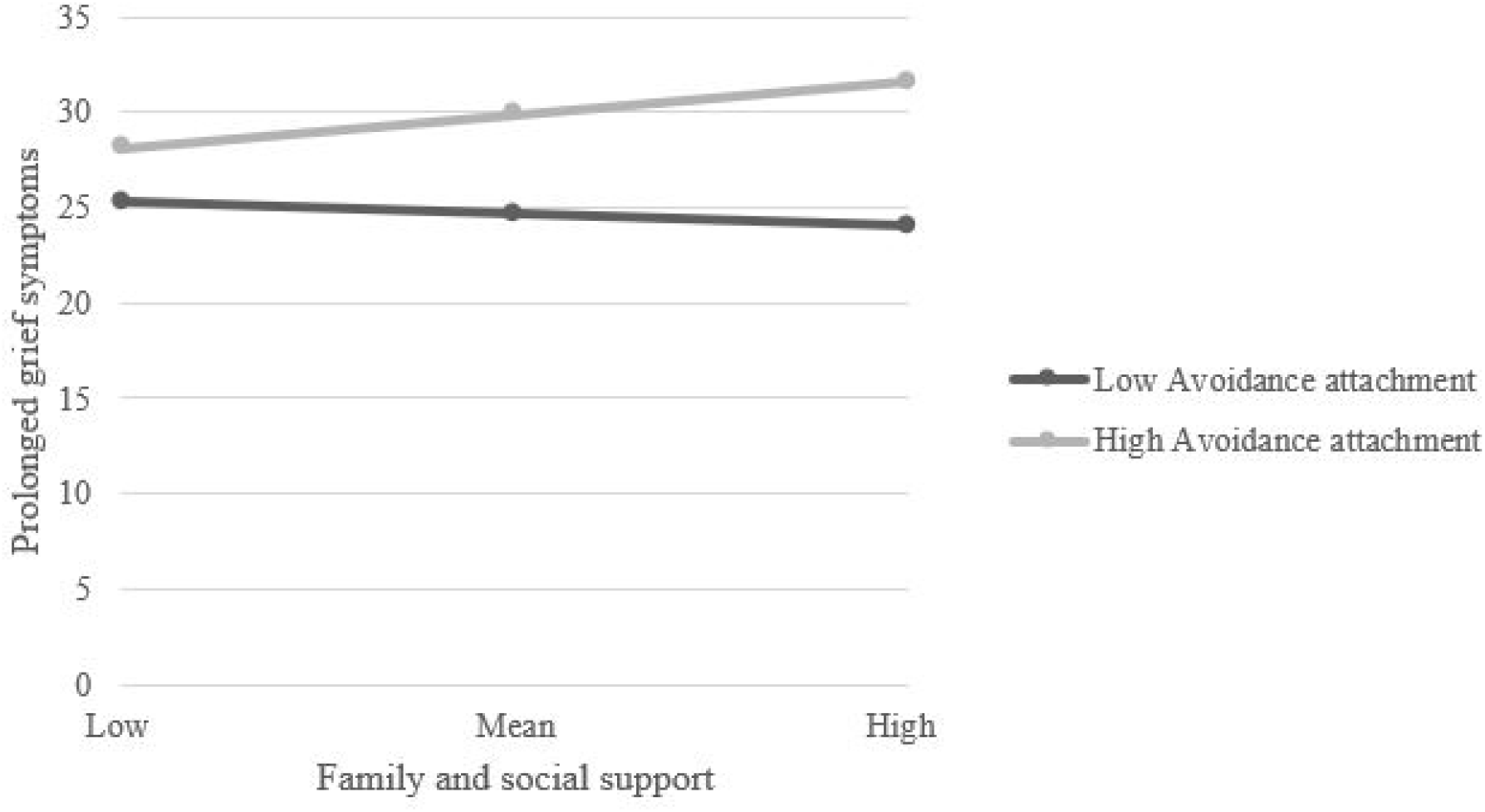
Interaction between perceived support and avoidance attachment for predicting prolonged grief symptoms among family caregivers

Table 4 shows that the Anxious attachment factor had a significant effect on the prolonged grief symptoms (*B* = 0.14; *p* < .01). As for the previous model, perceived support was not a significant predictor for prolonged grief symptoms. Furthermore, the interaction between the perceived support and the Anxious attachment factor was not significant.

**Table 4.**
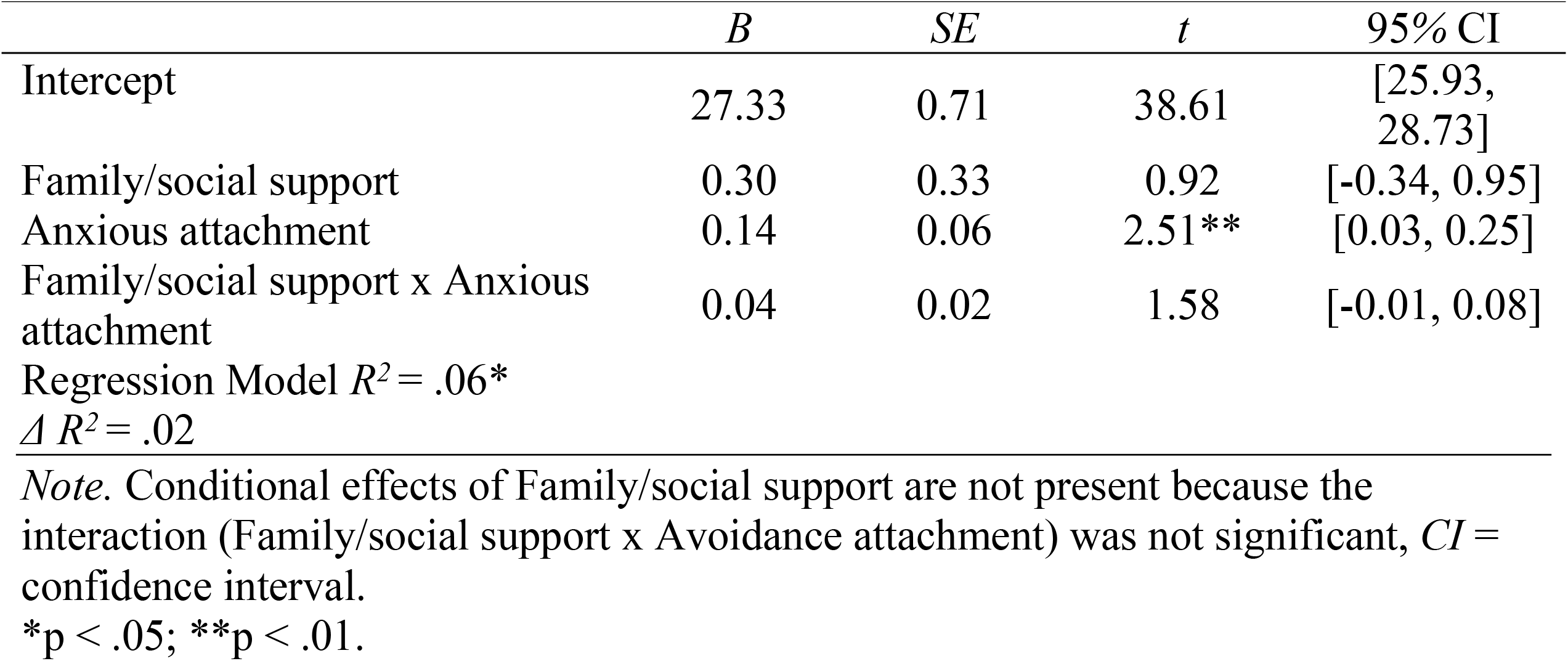
Anxious attachment moderates the relationship between Family/social support and intensity of prolonged grief symptoms

## Discussion

This study aimed to explore the interplay between the perceived support, insecure attachment factors, and the intensity of prolonged grief symptoms among caregivers who lost for cancer a family member. This general intent was addressed through two specific aims. First, we sought to examine the correlational coefficients between the family and social support during the assistance, avoidance and anxious attachment factors, and prolonged grief symptoms. As expected, we found a moderate and positive correlation coefficient between the two insecure attachment factors. This finding is coherent with previous research highlighting that avoidance attachment factor and anxious attachment factor are not incompatible [34]. Another finding of correlation analysis was that support from loved ones and friends was inversely associated with both factors of insecure attachment. When considering this relationship, findings from past studies showed analogous results, even though none of the studies have considered bereaved subjects in the context of palliative care [36]. Worth noting, we remind that attachment style during adulthood may influence the extent to which individual considers support from family and social in general. Thus, family caregivers of patients assisted in palliative home care with insecure attachment style may perceive lower support or may repute it as not important. Nonetheless, our interest was also to investigate the relationship between insecure attachment and symptoms of prolonged grief. Besides improving our understanding of the prevalence and characteristics related to the onset of prolonged grief disorder, findings from this field of research may be useful for its prevention. This should be considered in the context of palliative care as one path to strengthen the role of health psychology [8]. As revealed by our results, the avoidant but not anxious attachment factor was related to the reported symptom of grief. Since that Deutsch [37] has introduced the term “Absence of Grief” for understanding a lack of conscious affect following a loss, the avoidance attachment style has been proposed to explain fewer symptoms of prolonged grief in bereaved people, even though empirical research has produced contradictory results [25]. Whatever the cause of such inconsistent results, a promising line of research has investigated not only the relationship between attachment and grief but also the role of some moderating variables [38]. In this vein, the second aim of this was to verify if avoidance and anxious attachment factors impinge on the relationship between the perceived support and symptoms of prolonged grief in family caregivers. The burgeoning literature has well demonstrated that cancer patients who perceived low social support showed higher levels of depression and a worse quality of life [39-40]. However, as regards bereaved individuals, it is worth highlighting that the role of social support in protecting from adverse mental health outcomes remains ambiguous [12]. It has been argued that social support cannot sufficiently mitigate the loss of a loved one [41]. As depicted by our findings, there was not a direct effect of the perceived support on the intensity of prolonged grief symptoms among the bereaved people composing our sample. For the sake of clarity, it is worth emphasizing that the low reported support may be reveal conflicting relations, rather than the lack of supportive relationships [42], which in turn may be related to the functioning of the attachment system. Of particular interest in our study is that both avoidance and anxious attachment factors showed a direct effect on the reported symptoms of grief. Nonetheless, avoidance attachment seems to moderate the effect of perceived support on the outcome of bereavement. Put another way, how the support is perceived by the individual may hinge on its attachment style on the grounds that, in turn, it influences the extent to which he or she considers essential support from family members and friends. Turning to the two dimensions that characterize insecure attachment (i.e., avoidance and anxious ones), Mikulincer [25] argued that their balancing weight is paramount to coping with grief. Hence, taken together with results of correlational analysis, our findings may indicate an imbalance in this dynamic, even though more research focusing on bereaved individuals who lost a family member in the context of palliative care is needed. Indeed, knowledge on the factors increasing the risk of prolonged grief disorder in the context of palliative care would have practical value for its prevention.

Although our findings increase the understanding of the risk of prolonged grief disorder in the context of palliative care, there are also some limitations that future research should take into account. First, this study adopted a cross-sectional design that did not allow us to conclude without uncertainty for a causal relationship between the observed variables. Future studies adopting a longitudinal design would better account for the long-lasting role of insecure attachment style on the association between perceived support and prolonged grief symptoms. Second, the oversampling of certain characteristics (i.e., the female gender) among the participants may have influenced the results and whereby may be not generalizable to other bereaved individuals. Nonetheless, some characteristics of our sample such as a majority of female gender can be considered as representative of family members assisting their loved ones with a terminal illness.

## Conclusions

In sum, our results suggested that avoidance attachment behaves as a moderator of the relationship between perceived social support and symptoms of prolonged grief disorder among bereaved caregivers. Despite both avoidant and anxious attachment showed an effect on the intensity of prolonged grief symptoms, the interaction with perceived support was significant only for the avoidance attachment dimension. These results highlight the specificities among people with different insecurities attachment facing the loss of a loved one. Indeed, support from family members and friends may be helpful, even though high levels of avoidant attachment may interfere with the grief resolution and worsen the outcomes. Specialists and researchers within the field of palliative care may find useful our findings when assessing and preventing prolonged grief disorders among bereaved individuals.

## Data Availability

The data that support the findings of this study are available from the corresponding author, VL, upon reasonable request.

